# Outcome evaluation of effect of organizational strategies on reduction of patient waiting time at the outpatient department at Kilimanjaro Christian Medical Centre – Northern, Tanzania

**DOI:** 10.1101/2024.02.14.24302723

**Authors:** Henry Mollel, Manasseh Joel Mwanswila

## Abstract

**Background:** The Tanzanian healthcare system has long grappled with extended waiting times in outpatient departments (OPD). Studies at Kilimanjaro Christian Medical Centre (KCMC) revealed an average wait of six hours, marking KCMC with the longest waiting time among Tanzanian referral hospitals. Thus, this study was undertaken to assess the efficacy of the programme interventions and their effect on waiting time at KCMC, OPD.

**Methods:** An analytical cross-sectional approach quantitatively and qualitatively examined the subject. The study enrolled 412 patients who completed structured questionnaires, while 14 participants underwent in-depth interviews (ID) (8 healthcare providers, 6 patients) from 3^rd^ to 14^th^, 2023. Documentary review supplemented data. Quantitative analysis included descriptive statistics, bivariable, and multivariable techniques. Qualitative data underwent thematic analysis. Significance tests were at a 5% level.

**Result:** The overall OPD waiting time significantly decreased to 3.30 hours IQR (2.51-4.08) in contrast to the previous 6 hours prior to the intervention, showing the effectiveness of the intervention achieving a reduction of waiting time by 55%. Improvements were particularly evident, waiting time for registration (9 minutes), payments (10 minutes), triage (14 minutes for patients with insurance) and pharmacy (4 minutes). The implementation of Ushers emerged as a significant predictor to patient waiting time (AOR = 2.08, 95% CI, 1.10-3.94, p-value=0.025). Based on the IDI, the findings indicate a favourable change in patients’ attitudes towards waiting time at the OPD. However, there is skepticism regarding the expansion of hospital infrastructure and its effect on waiting time, as well as reliance on auxiliary support due to insufficient human resources.

**Conclusion:** Even though the established intervention strategies have managed to reduce waiting time, additional measures to attain the global standard of waiting time from 30 minutes to 2 hours are needed.

## Introduction

The healthcare system in Tanzania has been facing a lot of challenges with prolonged waiting times in the hospital outpatient department (OPD). The reported contributing factors include the increased need for healthcare due to uncontrolled population growth, inadequate medical experts, underdeveloped healthcare systems, and ineffective referral systems [1]. The audit report from the Ministry of Health on the management of referral and emergency healthcare services at zonal and regional referral hospitals showed high OPD waiting time. Previous studies further suggest that the average waiting time at, Muhimbili National Hospital OPD was 4 – 6 hours; Muloganzila Zonal Referral Hospital was 3 – 4 hours; Bugando Medical Centre was 2.5 hours, Mbeya Zonal Hospital was 3 – 4 hours and Kilimanjaro Christian Medical Centre (KCMC) was 6 hours [1,2]. This data suggests that KCMC has the longest waiting time of all the Zonal and National Referral Hospitals in Tanzania. In response to this long waiting time KCMC implemented a programme of interventions to address the outpatient waiting time in which were integrated into the strategic plan spanning from 2016 to 2021. The interventions included expansion of hospital infrastructure, streamlining patient flow system, deployment of ushers, and the additional of human resources.

Expansion of hospital infrastructure is essential to accommodate the growing demand of healthcare services. Studies conducted by researchers have highlighted the pressing need for improved facilities and consequences of inadequate infrastructure on patient experiences. [3] conducted an institutional based observational descriptive study with a cross-sectional design which involved 100 patients seeking outpatient services at Apollo Hospital Medical College, in Hyderabad, India. The aim of the study was to assess the time taken at different service delivery points in the outpatient department and to assess the perception of beneficiaries regarding the total time spent in the OPD. The results revealed that the number of patients requesting OPD services had multiplied, but OPD facilities had not kept pace with this growth. Diri & Eledo, (2020) conducted a comparative descriptive survey among 300 patients visiting three major public hospitals in Nigeria in Yenegoa, Bayelsa state. The study aimed at investigating and comparing the performance of health workers in delivering various services in the reduction of patients’ waiting time. The patients who attended the OPD raised a concern that the OPD has got inadequate space to accommodate the large number of patients, and it needed to be enlarged. Sarwat, (2022) conducted a cross-sectional study among 402 patients visiting a tertiary care hospital in Pakistan. The study aim was to assess patients’ satisfaction, factors, and effects of waiting time. The results ascertained that factors contributing to delayed waiting time included insufficient examination rooms and poor physical layout of the OPD.

The effective management of patient flow and waiting time within the OPD remains a critical area of concern. One promising strategy to optimize patient experiences and streamline operations involves the deployment of ushers or stewards within the OPD. Deployment of ushers or stewards within the OPD has brought in attention as a proactive strategy to address the challenges associated with patient. Yadav (2017), conducted an observational descriptive study with institutional based cross-sectional design which involved 100 patients seeking outpatient services at Apollo Hospital Medical College, Hyderabad in India. The aim of the study was to assess various issues related to patient waiting time in the OPD. The study found a significant reduction in waiting was achieved at the OPD after additional members of staff were appointed as patient care coordinators to guide patients. An observational study done in Haiti to identify the factors that contribute to lengthy wait times for health services and methods to shorten them, found that patients lacked on navigating the hospital. This was a contributor to long waiting times and justified the importance of having ushers placed within the OPD [6]. In Ethiopia, a cross-sectional study was conducted by Geta & Edessa, (2020) at the OPD with a sample size of 420 patients. The aim of the study was to assess patient satisfaction with waiting time among outpatients and associated factors at Nekemte referral hospital. The findings from the study revealed that 37.3%, (n=119) of participants commented that there were no directional signs on how to navigate well within the OPD hence prolonging their waiting time. Chandra (2017), conducted a systematic survey which aimed at reducing waiting time of OPD patients in hospitals using different types of models. Based on the findings it was suggested that ushers should be appointed in order to guide patients within the OPD and by doing so time will not be wasted. Carter et al., (2015) conducted a pilot study evaluating the impact of introducing patient navigators, who act as ushers or stewards, on reducing patient waiting time among 100 patients attending a tertiary OPD in the USA. The study found that patient navigators significantly reduced patient waiting times and improved patient satisfaction. Patient navigators were able to identify patients who needed assistance and guide them through the care process, which helped to streamline patient flow and reduce waiting times. Additionally, patients felt more supported and informed throughout their visit, which led to higher levels of satisfaction.

Mukherjee et al., (2017) conducted a quasi-experimental design with pre and post intervention comparison groups among 343 patients with 179 patients in the pre-intervention group and 164 patients in the post-intervention group. The study aimed to evaluate the impact of introducing ushers as an intervention to reduce patient waiting times in an outpatient clinic in the United Kingdom. The ushers were responsible for greeting patients, providing information about wait times, and directing patients to the appropriate clinic areas. The study found that the introduction of ushers was associated with a significant reduction in patient waiting times, from an average of 35 minutes before the intervention to 18 minutes after the intervention. The authors attributed this reduction to the improved patient flow and decreased patient anxiety resulting from the ushers’ clear communication and direction. The authors also noted that the introduction of ushers had a positive impact on staff satisfaction, as staff felt that they were better able to manage patient flow and improve the overall patient experience. Overall, this study provided evidence for the potential effectiveness of introducing ushers or stewards as an intervention to reduce patient waiting times in a tertiary OPD in a developed country.

Chakravarty et al., (2016) conducted a non-randomized controlled study with pre and post intervention design. The sample size for the study was 1,500 patients, of which 750 were enrolled in the intervention group and 750 were enrolled in the control group. The intervention involved the introduction of hospital ushers who were responsible for assisting patients and their families with various tasks during their visit, such as registration, wayfinding, and providing information. The aim of the study was to evaluate the impact of introducing hospital ambassadors, similar to ushers or stewards, on patient satisfaction and waiting times at a tertiary care hospital in India. The study found that the introduction of hospital ambassadors led to a significant improvement in patient satisfaction and reduction in waiting times. Patient satisfaction improved from 70% to 88% after the introduction of hospital ambassadors. Additionally, the average waiting time for patients decreased from 43 minutes to 20 minutes.

In Pakistan, Sarwat, (2022) conducted a cross-sectional study among 402 patients visiting a tertiary care hospital in Pakistan. The study aim was to assess patients’ satisfaction, factors, and effects of waiting time. The findings revealed that one of the factors of delayed waiting time to patients was lack of guidance on how to navigate within the OPD.

The insufficiency of healthcare professional, especially in low resource settings remains a global concern, significantly affecting patient care and waiting time. Studies have consistently identified the shortage of healthcare staff as factor contributing to prolonged waiting time within the OPD. The World Health Organization (WHO) approximates that in 90% of low-income countries, there is a significant shortage of healthcare professionals, defined by having fewer than 4.4 qualified staff members per 1000 individuals [12]. A study conducted by Safdar et al., (2020) aimed to create a model for assessing queues to analyse the influx of walk-in outpatients in a busy public hospital in Pakistan. The researchers employed Data Envelopment Analysis (DEA) to construct a composite Queuing-DEA model for evaluating patient wait times. The results indicated that a shortage of healthcare staff emerged as a crucial factor contributing to extended waiting time in the outpatient department.

Adamu & Oche (2013) looked into factors influencing patient waiting times in the GOPD of a tertiary healthcare facility located in northern Nigeria. This descriptive cross-sectional investigation involved a sample of 100 patients. The study results indicated that the most prevalent cause of long waiting times in the GOPD was the substantial patient load relative to the limited healthcare workers. As a conclusion, the study emphasized the critical necessity for augmenting the healthcare workforce within GOPDs to address this issue effectively. In Kenya, a cross-sectional study design was conducted by Wafula & Ayah, ( 2021) among 384 patients attending the staff clinic at the University of Nairobi Health Services to assess waiting times and associated factors. The findings indicated that a majority of respondents (52%) believed that enhancing staff availability at their respective stations would contribute to reduced patient waiting times. It was concluded that the primary factor contributing to prolonged waiting times was the insufficient number of healthcare providers. Amani et al., (2021) conducted a qualitative study aimed at exploring the perspectives and experiences of healthcare services among elderly individuals, both insured and uninsured, residing in rural Tanzania. The study employed an exploratory qualitative design which used eight focus group discussions, and involved a purposive selection of 78 elderly participants of both genders. The findings identified a scarcity of healthcare professionals resulting in prolonged waiting times and restricted consultation durations with physicians, ultimately affecting the overall quality of care.

In Tanzania, the Ministry of Health has reported a notable rise in the number of healthcare workers, rising from 29,063 in 2006/07 to 102,919 in 2019. Despite this increase, there remains a significant shortfall in human resources for health (HRH). Currently, this shortage is estimated to be at 52 percent of the actual need. Although the ratio of health professionals per 10,000 people has seen an increase from 15.7 in 2010 to 17.2 in 2020, the scarcity of personnel remains a pressing concern [17]

In Tanzania the Ministry of Health has not established the gold standard waiting time for patients to wait for services at the OPD [2]. However the United States Institute of Medicine (IOM) has established their gold standard patient waiting time at the OPD which suggests that medical care should be provided to at least 90% of patients no later than 30 minutes after their scheduled appointment time [18,19]. The Patient’s Charter of UK, has recommended the same standard as the IOM (Diri, and Eledo, 2020; Lee et al., 2022). The absence of a gold standard waiting time carries several significant implications. It results in inconsistent patient experiences with unpredictable waiting times across facilities, leading to frustration and dissatisfaction. Prolonged and varied waiting times can compromise the quality of care, affecting patient outcomes. Inefficient resource allocation becomes a challenge, hampering the ability to determine staffing and infrastructure needs [21]. This lack of a benchmark reduces accountability, and healthcare providers may not be incentivized to improve waiting times. It adversely affects patient satisfaction, the reputation of healthcare providers, and can exacerbate healthcare disparities [5].

Therefore, the aim of this study was to assess the effectiveness of organizational strategy interventions implemented at KCMC from 2016 to 2022 in reducing patient waiting times at the OPD. Additionally, the study aimed to address the lack of information regarding the current waiting time status influenced by these interventions. In conclusion, the study successfully achieved its aim.

## Materials and methods

### Study design and setting

This was an outcome evaluation which employed analytical cross-sectional study which was part of a lager mixed method study conducted consecutively for two weeks from 3^rd^ to 14^th^ July, 2023. The study was conducted at the KCMC Outpatient Department. This area was chosen because the outpatient department at Kilimanjaro Christian Medical Centre sees high volume of patients on regular basis from diverse backgrounds, including rural and urban populations of Tanzania as well as neighbouring countries. For instance in the year 2022, a total of 277,013 (92%) attended the OPD. This could potentially result in longer waiting time for patients at the OPD, making it a suitable location for studying patient waiting time.

### Sampling and sample size

The study surveyed 412 and 14 patients both quantitatively and qualitatively respectively. The quantitative sample size was obtained using Cochran, (1977) formula:

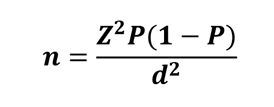

Whereby: -

n = sample size

Z = is the standard normal deviation which is 1.96 for 95% confidence interval

P = is the percentage of patients attending the OPD at KCMC is estimated to be 0.5, attributed to the absence of prior research data

d = is the margin of error, which is 5% (0.05)

Therefore,

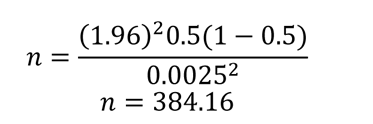

Therefore, calculate minimum sample size of this side was 384 patients approximated to be 422 after adjustment of 10 percent non response rate.

While on qualitative sample size, this research adopted the sample size of 12 respondents in which Boddy, (2016), suggested that with practical research indicating that data saturation in a relatively homogeneous population could occur in 12 sample sizes. The study focused on patients aged 18 and older who attended the OPD during the data collection period. In addition, patients who were severely ill, had scheduled admission appointments were excluded as well as first time attendees (new patients) were excluded because they lacked prior experience with the implemented interventions. Additionally, healthcare providers at the OPD were interviewed to assess the effect of intervention strategies on waiting time. Further, in terms of the sampling method, a convenience sampling method was used to include all patients who were present at the OPD during data collection, chosen for its accessibility in engaging study participants, while healthcare providers in the OPD were selected through purposive sampling.

### Data collection tool and procedure

In this study, the researcher employed a modified structured questionnaire as a data collection tool for gathering information from the patients as inspired by Sundresh & Nagmothe’s, (2017) study. The tool had socio-demographic characteristics which included age, gender, marital status, education level, occupation, place of address, mode of payment & Year of attendance at KCMC. The measurement scale for organizational strategy was typically ordinal, based on eight (8) likert scale questions with response options of 1=strongly disagree, 2=disagree, 3=neutral, 4=agree and 5=strongly agree. Allowing patients to indicate their level of agreement or disagreement with statements related to organizational strategies. Additionally, strongly disagree and disagree were consolidated as disagree and neutral, agree and strongly agree were consolidated as agree following the approach used in a previous by study [25]. The internal reliability of the eight items used to assess effectiveness of organizational strategies on reducing patient waiting time was measured using Cronbach’s alpha = 0.908. The survey included questions on arrival time, time queue number was issued, registration waiting time, payment waiting time, triage waiting time, waiting time to see the doctor, pharmacy waiting time, laboratory waiting time, radiology waiting time and exit time. These data were collected from patients who attended clinics such as the OPD clinic, orthopedic clinic, Medical clinic, surgical clinic, Urology clinic, Ear, Nose & Throat, Diabetic, cardiac clinic, Neurology and Neurosurgery. Waiting time was measured with a stopwatch.

Semi-structured interview guide for conducting in-depth interviews with patients and healthcare providers was developed and the interview guide had the questions on the following, socio-demographic characteristic such as gender, age, marital status, education level, occupation, and address, and questions on organizational strategies such as expansion of hospital infrastructure, patient flow, presence of ushers and additional human resources.

Also, the researcher conducted a documentary review, analyzing written records detailing time allocation before the studied event. This approach offered insights into past practices, aiding pattern and trend analysis. It involved reviewing benchmarks like a six-hour average waiting time, median wait time for specific clinics, and total treatment duration for patients in various clinics.

### Variables and measurements

#### Dependent variable

The study defined the dependent variable as follows: overall patient waiting time, which was captured using a stopwatch, was categorized as a binary dummy variable. A value of 1 represented OPD waiting times less than 3 hours, while a value of 0 indicated OPD waiting time exceeding 3 hours. Comparison with Standards: The analysis involved evaluating OPD waiting time against established benchmarks. This included comparing the waiting time with the standards outlined in the Patients Charter of the United Kingdom (UK) and the recommendations from the United States Institute of Medicine (IOM), which advocate that at least 90% of patients should receive medical care within 30 minutes of their scheduled appointment time. Additionally, the study compared the observed 6-hour waiting time, set as outpatient waiting time at KCMC Zonal Hospital, to assess whether there was any reduction post-intervention.

#### Independent variables

Independent variables included gender (male or female), education, mode of payment (cash or insurance), sufficient examination rooms, patient flow system, deployment of ushers, guidance by ushers, additional human resource, expansion of hospital infrastructure, OPD space, allocation of medical records personnel and cashier within clinic premises.

Statistical Tests: To explore potential associations between dependent and independent variables, statistical tests were employed. Logistic regression analysis, encompassing both bivariate and multivariate analyses, was conducted. The multivariable analysis included all variables with p<0.200 as identified during the bivariable analysis. It was further adjusted for gender, level of education, and mode of payment. All statistical analyses were conducted at a significance level of 0.05. These analytical steps were taken to provide a comprehensive evaluation of the effect of the organizational strategies intervention on patient waiting times.

### Data analysis

#### For quantitative approach

The data collected were then imported to the STATA 18.0 for further analysis. Descriptive Statistics: The analysis began with the presentation of data using various methods, including figures, graphs, and frequency distributions. Further, to determine the effect each response was rated on a scale of 1 to 5. The calculation involved subtracting the average from the total number of respondents. This process was performed for the questionnaires focusing on organizational strategies. Subsequently, cut-off points were utilized for each area to categorize the effectiveness of each intervention strategies as follows: 1-1.8 (very low), 1.8-2.6 (low), 2.6-3.4 (medium), 3.4-4.2 (high), and 4.2-5 (very high) [26]. Also, in this study, efficacy was determined by calculating the percentage reduction in OPD waiting time achieved through the implementation of intervention strategies. This calculation involved using the current overall OPD waiting time (as shown in Table 4) as the numerator and the 6-hour benchmark as the denominator [2].

#### For qualitative approach

All interview transcripts were transcribed verbatim and translated into English language. In order to maintain the original meaning back translation was employed. The analysis was done using the English transcript. Thematic data analysis was employed using both deductive and inductive reasoning. Consequently, a preliminary codebook for data analysis was developed, aligning with the study objectives, after which the final codebook was imported into Atlas.ti 7.0 qualitative data analysis computer software. Inductive coding was assigned to text segments which built on emerged new themes that were not pre-determined. The codes were sorted into categories then were clustered into sub-themes which were aligned into themes. The entire process of analysis was iterative.

#### Ethical clearance

Ethical Clearance Committee from Mzumbe University from the Directorates of Research, Publication and Postgraduate provided ethical clearance with reference number MU/DPGS/INT/38/Vol. IV/236. Subsequently, the proposal was submitted for evaluation to the College Research Ethics and Review Committee (CRERC) at Kilimanjaro Christian Medical University College – Moshi. The CRERC granted approval, as indicated by certificate number 2639. Additionally, the data collection procedure received endorsement from the directors of KCMC Hospital reference number KCMC/P.1/Vol. XII. Prior to data collection, participants provided written informed consent. We explained the research objectives to ensure transparency and maintained confidentiality.

## Results

### Social demographic characteristics of the study population

In this study, the initial calculated sample size was 422 patients. However, out of this group, only 412 patients consented to participate and completed the questionnaire. This resulted in a response rate of 97.6%. The median age was 52 (IQR, 38-65), with a majority aged over sixty. Over half were female (53.6%, n=221), and the majority were married (76%, n=313). Most had basic education, including primary (44.7%, n=184) and secondary education (26.7%, n=110). More than half were peasant farmers (52.4%, n=218), and a vast majority (94.7%, n=338) resided within the KCMC catchment area. The majority were insurance patients (82.0%, n=338), and more than two-thirds (66.5%, n=274) had attended KCMC before the intervention’s inception (Table 1).

**Table 1:** Social demographics characteristics of patients for quantitative.

| Variable | n | Percentage (%) |
| --- | --- | --- |
| **Gender** |  |  |
| Male | 191 | 46.4 |
| Female | 221 | 53.6 |
| **Age-group** |  |  |
| 18 – 35 | 89 | 21.6 |
| 36 – 45 | 70 | 17.0 |
| 46 – 60 | 114 | 27.7 |
| 61+ | 139 | 33.7 |
| **Age (Median, [IQR])** | 52, [38 - 65] |  |
| **Marital status** |  |  |
| Married | 313 | 76.0 |
| Single | 64 | 15.5 |
| Divorce/Separated | 4 | 1.0 |
| Widow/widower | 31 | 7.5 |
| **Level of education** |  |  |
| Not attended school | 20 | 4.9 |
| Primary | 184 | 44.7 |
| Secondary | 110 | 26.7 |
| College | 61 | 14.8 |
| University | 37 | 9.0 |
| **Profession/Care** |  |  |
| Peasant farmer | 218 | 52.4 |
| Business | 80 | 19.4 |
| Employed | 86 | 20.9 |
| Self-employed | 20 | 4.9 |
| Student | 10 | 2.4 |
| **Residence** |  |  |
| Within KCMC Catchment area | 390 | 94.7 |
| Outside KCMC Catchment area | 22 | 5.3 |
| **Mode of payment** |  |  |
| Cash | 70 | 17.0 |
| Insurance | 338 | 82.0 |
| Exempted | 4 | 1.0 |
| **First Year of attendance at KCMC** |  |  |
| 2018 and below | 164 | 39.8 |
| 2019 | 34 | 8.3 |
| 2020 | 33 | 8.0 |
| 2021 | 43 | 10.4 |
| 2022 | 54 | 13.1 |
| 2023 | 84 | 20.4 |
| **Patient attendance at KCMC** |  |  |
| After intervention | 138 | 33.5 |
| Before intervention | 274 | 66.5 |

### Demographic characteristics of qualitative sample

A total of 14 participants comprising of 8 healthcare providers and 6 patients were enrolled. More than half (57.1% n=8) of participants were males (Table 2).

**Table 2:** Demographic characteristics for qualitative sample.

| Variable | n | Percentage (%) |
| --- | --- | --- |
| **Gender** |  |  |
| Male | 8 | 57.1 |
| Female | 6 | 42.9 |
| **Age-group** |  |  |
| 36 – 45 | 2 | 14.3 |
| 46 – 60 | 8 | 57.1 |
| 61 and above | 4 | 28.6 |
| **Marital status** |  |  |
| Married | 14 | 100 |
| **Level of education** |  |  |
| Primary | 1 | 7.1 |
| Secondary | 1 | 7.1 |
| College | 6 | 42.9 |
| University | 6 | 42.9 |
| **Profession/Care** |  |  |
| Employed | 12 | 85.7 |
| Self-employed | 2 | 14.3 |
| **Residence** |  |  |
| Within KCMC Catchment area | 14 | 100 |

### Themes emerged from IDIs

The sub-themes identified from the findings encompass various aspects of the OPD functioning and their impact on patient waiting time. Such as improved efficiency at the reception, challenges with doctor availability, technological and infrastructure enhancement, limited space, dual use of facilities, improved overall OPD waiting time, skepticism regarding effect on waiting time, enhanced patient workflow and reliance on auxiliary support (Table 3).

**Table 3:** Themes emerged from IDIs.

| Theme | Sub-theme |
| --- | --- |
| Registration area | Improved efficiency at the reception |
| Waiting to see the doctor | Challenges with doctor punctuality |
| Pharmacy area | Technological and infrastructure enhancement |
| Laboratory area | Limited space |
| Radiology area | Challenge with dual use of same facilities |
| OPD waiting time | Improved overall OPD waiting time |
| Expansion of hospital infrastructure | Skepticism regarding effect on waiting time |
| Deployment of Ushers | Enhanced patient workflow |
| Additional human resource | Reliance on Auxiliary support |

### OPD waiting time since the inception of implementation of the interventions

The objective of this study was to assess waiting times for registration, payment, triage, doctor consultations, pharmacy services, laboratory procedures, radiology tests, and calculate the overall waiting time in the OPD.

Following the intervention, it was observed that the overall median waiting time in the OPD was reduced to 3.30 hours IQR (2.51-4.08) in contrast to the previous six-hour (6) waiting time prior to the intervention. Specifically, the median waiting time for registration was 9 minutes IOR (0.03-0.15). For payment, the median waiting time was 10 minutes IOR (0.07-0.15). For triage patients using out-of-pocket payments experienced median waiting time of 17 minutes IQR (0.05-0.19) while those with insurance had a slightly shorter median waiting time of 14 minutes IQR (0.06-0.19) and the median waiting time to see a doctor was 1.36 hours IQR (0.51-2.01). The time from arrival to actually seeing a doctor was measured at 3.08 hours IQR (2.13-3.30). Furthermore, the median consultation time was 19 minutes IQR (0.15-0.24), median waiting time at the pharmacy was 4 minutes IQR (0.02-0.06), at the laboratory it was 31 minutes IQR (0.20-0.37) and waiting times at Radiology varied based on the specific service. X-ray services in different rooms had median waiting times ranging from 35 minutes to 1.15 hours with varying IQR (0.23-2.19). Ultrasound services had median waiting time of 32 minutes (Table 4).

### Qualitative findings Registration area

#### Improved efficiency at the reception

Following the adoption of electronic medical records (EMRs) it has undeniably enhanced the overall efficiency of our registration process, benefiting both patients and staff. This was testified by a male patient

> *“I have been receiving treatment here at KCMC for over 20 years. In the past, here at the reception of the medical records department, it was necessary to have someone, a staff member, whom you would contact in advance, preferably three days before your clinic day, so that they could start looking for your file. This way, you could save time waiting. However, nowadays, this process is no longer in place. When I arrive, I simply present my card, and in no time, I’m on my way to the next area. There’s no longer any time wasted at the reception.”* (IDI – Male Patient)

Another interviewee added that:

> *“Nowadays, with the system in place, the process is streamlined, allowing me to efficiently register as many patients as possible in a short amount of time. I no longer have to leave the reception area to search for files, which has significantly improved the efficiency of the registration process.”* (IDI – Male healthcare provider (HCP))

### Waiting time to see the doctor

#### Challenges with doctor punctuality

The issue of waiting times for patients to see the doctor has emerged as a significant concern within the healthcare facility. This concern is consistently echoed in both the quantitative data and qualitative interviews.

For example a female HCP had to say this:

> *“*[…] *commencing clinics promptly can be challenging for doctors, as it is crucial for them to first participate in the morning report, which provides essential updates on the status of hospitalized patients”* (IDI – male HCP).

After probing as to why they cannot split two teams of doctors so that one team should attend to outpatients:

> *“We have a limited number of doctors, making it challenging to divide them into two groups. Moreover, admitted patients demand our additional attention, as some rely on oxygen for breathing, while others are too ill to walk. Unlike outpatients, the majority of whom can independently come for treatment, we kindly request their understanding as we prioritize the care of our admitted patients”* (IDI – male HCP).

A female patient gave some observations

> *“[….] Mmh! I want to highlight that delay in seeing the doctor can have serious consequences. It can lead to a worsening of symptoms or conditions, increase stress levels, and ultimately result in reduced satisfaction with the healthcare service. It’s imperative that we address these extended waiting times. This is crucial not just for the comfort of the patient, but also to ensure that medical care is administered in a timely and effective manner”* (IDI – female patient).

### Pharmacy

#### Technological and infrastructure enhancement

In the pharmacy department, there has been a notable improvement in waiting times. Patients now experience a comfortable and efficient process, with minimal time spent before receiving their prescribed medications. This sentiment was echoed by both patients and healthcare providers who were interviewed who said that: -

> *“With the use of a computerized system, things have been greatly simplified. The waiting time to collect medicine has become short. When I come here, I wait for just a little while and quickly get my medicine.”* (IDI – Male patient).

> *“Apart from using the computerized system in place, which has simplified things, the hospital administration has managed to establish three additional pharmacies apart from this one, thus reducing congestion in a single pharmacy, as it used to be in the past. That’s why now a patient can be served quickly”* (IDI – male HCP).

A female patient also shared her positive perspective on the improvements in waiting time at the pharmacy compared to the past. The interviewee emphasized that: -

> *“…. I have noticed a significant improvement in the waiting time at the pharmacy. Previously, it used to take much longer causing inconvenience and frustration. However with the recent changes and enhancements in the system, the waiting time has been noticeably reduced. Now, I spend considerably less time waiting to collect my medications, which has made overall experience much more efficient and pleasant. It’s a welcome improvement that has positively impacted the patient experience” (IDI – female patient)*.

### Laboratory department

#### Limited space

In the laboratory department, the waiting time has been a subject of varying experiences among patients. Some patients have reported relatively short waiting periods, while others have encountered longer durations.

> *“I have been patiently waiting for a long time to be called for my tests, not yet up to now”* (IDI – female patient).

Another interviewee shared that:

> *“I’ve noticed that one of the main reasons for long waiting times at the laboratory here is the limited space. The laboratory rooms at the Outpatient Department (OPD) have remained the same since the hospital was established, which means they can only accommodate a small number of patients at a time. This often leads to a backlog of patients waiting to get their tests done. It’s clear that expanding the laboratory facilities is crucial to reduce these extended waiting times and ensure more efficient service delivery for everyone”* (IDI – male HCP).

### Radiology department

#### Challenge with dual use of same facilities

Despite having modern diagnostic equipment, which has significantly contributed to reducing patient waiting times, there are still instances where patients experience long waiting time in the radiology department:

> *“For me, even though waiting for an X-ray may take some time, I don’t mind the wait. I’ve noticed a significant improvement in waiting times compared to before. In addition nowadays, when I have an X-ray, I can also consult with my doctor on the same day, which wasn’t possible in the past”* (IDI – male patient).

One interviewee highlighted a crucial factor contributing to the extended waiting times at the radiology department and pointed out that:

> *“The same rooms at the radiology department are utilized for both outpatient and inpatient cases. As a result, priority is often given to the admitted patients, leading to longer waiting times for those seeking outpatient radiology services. This dual-use of facilities poses a challenge in managing patient flow and significantly contributes to the observed delays in the radiology department”.* (IDI – male HCP)

### Patient OPD waiting time with Six (6) and Three (3) Hours Threshold

Not a single patient managed to complete the treatment within the recommended 30-minute window following their scheduled appointment. When assessed based on the KCMC benchmark of a 6-hour timeframe, the vast majority of patients (98.8%, n=407, 95% CI, 97.0%-99.5%) indicated that they received the OPD services within a period of less than six hours. However, when the time threshold was further reduced to three hours, 31% (n=128, 95% CI, 26.6%-35.6%) of all surveyed patients reported that they received OPD services within a duration of fewer than three hours (Figure 1).

**Figure 1:**
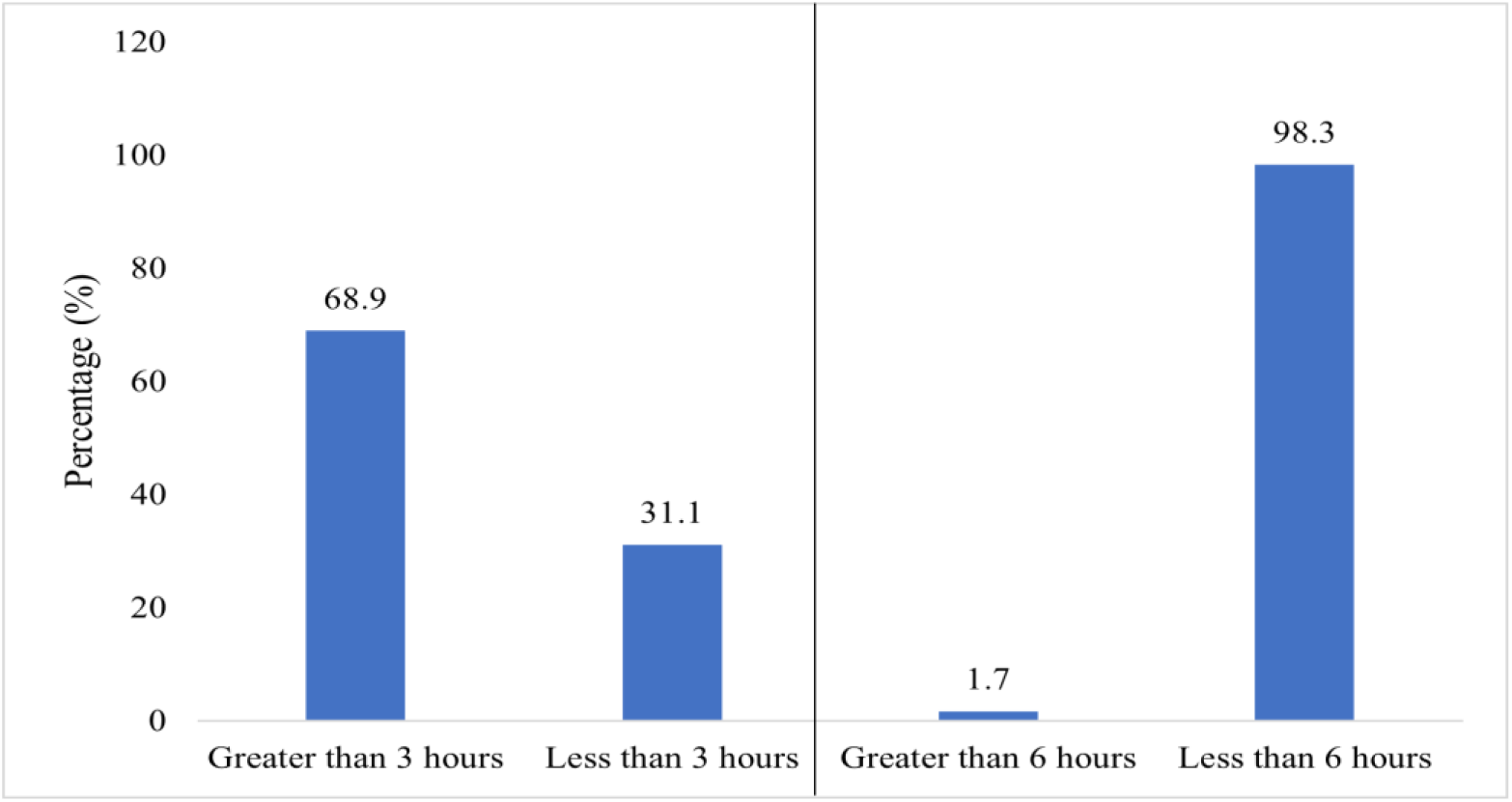
Patient OPD waiting time with six (6) and three (3) hours threshold (n=412) Qualitative findings.

### Improved overall OPD waiting time

Furthermore, during the in-depth interviews (IDIs), patients emphasized receiving OPD services within a timeframe of below three hours.

For instance, a male patient remarked:

> “*Certainly, drawing from my extensive experience of years attending KCMC hospital, I can attest to the positive changes in the waiting times for OPD services. Patients, including myself, are genuinely appreciative of this effective reduction in waiting times. I personally find it remarkable that I can now complete all the necessary OPD services in just about three hours, which is a stark contrast to the longer waiting periods we used to endure. This improvement has undoubtedly enhanced the overall patient experience and contributes positively to our healthcare journey”* (IDI – male patient).

### Effect of organizational strategies on patient waiting time

The study aimed to assess how organizational strategies influence the reduction of patient waiting time. These strategies were categorized into four domains, including the expansion of hospital infrastructure, patient flow system, deployment of ushers in the OPD, and addition of human resources. The data obtained through self-reporting were summarized descriptively using mean scores and standard deviations. The Likert scale ranging from 1 (strongly disagree) to 5 (strongly agree) was employed for measurement. Additionally, specific cutoff points were used to categorize factors contributing to the reduction of patient waiting times: 1-1.8 indicating very low effect, 1.8-2.6 denoting low effect, 2.6-3.4 representing a medium effect, 3.4-4.2 indicating a high effect, and 4.2-5 reflecting a very high effect.

Consequently, the overall average effectiveness of these organizational strategies was rated fairly high with an average score of 3.94 (SD=1.070). The expansion of infrastructure received a mean score of 3.88 (SD=1.088). Patient flow system had a mean score of 4.06 (0.974). Presence of Ushers in the OPD received a mean score of 3.99 (1.074). Additionally, the additional of human resources at the OPD had a mean score of 3.83 (SD=1.203). Table 5

**Table 4:**
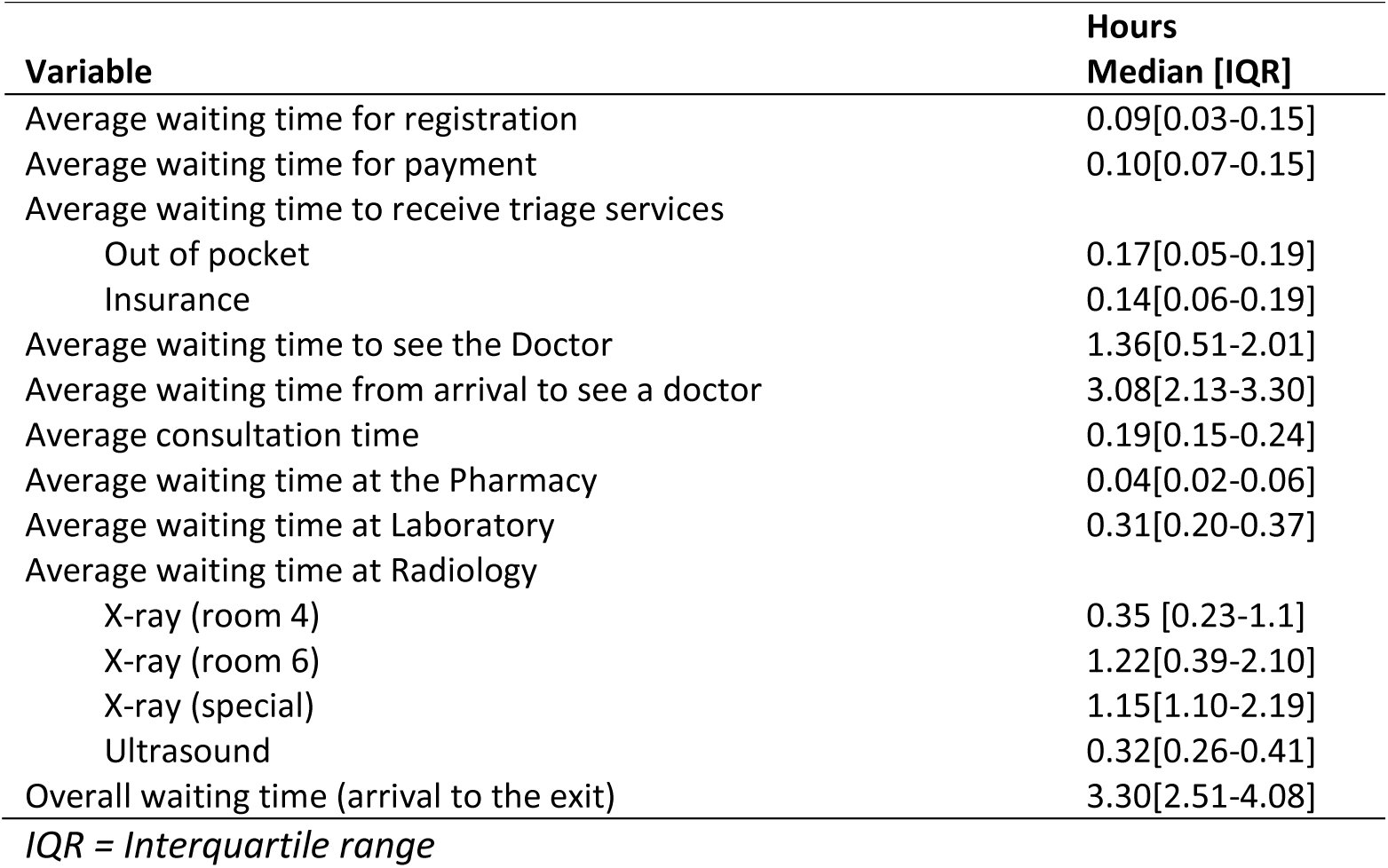
Distribution of Patient waiting time according at service areas.

**Table 5:** Organizational Strategies on Patient Waiting Time.

| Statement | Level of agreement |  |  |  |  | Mean (+SD) |
| --- | --- | --- | --- | --- | --- | --- |
|  | SD<br>N (%) | D<br>N (%) | N<br>N (%) | A<br>N (%) | SA<br>N (%) |  |
| **Expansion of infrastructure** |  |  |  |  |  |  |
| The OPD size has got enough space to accommodate the large number of patients | 12(2.9) | 26(6.3) | 21(5.1) | 210(51.0) | 143(34.7) | 4.08(0.952) |
| Expanding hospital infrastructure has reduced waiting time | 15(3.6) | 44(10.7) | 19(4.6) | 194(47.0) | 140(34.0) | 3.97(1.069) |
| There is sufficient examination rooms | 24(5.8) | 88(21.4) | 33(8.0) | 159(38.6) | 108(26.2) | 3.58(1.244) |
| **Overall mean** |  |  |  |  |  | **3.88(1.088)** |
| **Patient flow system** |  |  |  |  |  |  |
| Allocating medical records person and cashier within the clinic premises reduces waiting time | 9(2.2) | 19(4.6) | 19(4.6) | 193(46.8) | 172(41.6) | 4.21(0.895) |
| Streamlining patient flow system reduces waiting time | 14(3.4) | 46(11.2) | 27(6.6) | 203(49.3) | 122(29.6) | 3.91(1.052) |
| **Overall mean** |  |  |  |  |  | **4.06(0.974)** |
| **Presence of ushers** |  |  |  |  |  |  |
| Guidance by ushers on how to navigate within the OPD has helped to reduce time/ ushers' clear communication and direction | 12(2.9) | 23(5.6) | 18(4.4) | 203(49.3) | 156(37.9) | 4.14(0.944) |
| Presence of ushers/stewards has helped to reduce waiting time | 14(3.4) | 78(18.9) | 19(4.6) | 152(36.9) | 149(36.2) | 3.83(1.203) |
| **Overall mean** |  |  |  |  |  | **3.99(1.074)** |
| **Additional of human resource** |  |  |  |  |  |  |
| Additional human resource at the OPD has helped to reduce waiting time | 14(3.4) | 77(18.7) | 24(5.8) | 148(35.9) | 149(36.2) | 3.83(1.203) |
| **Grand Overall mean** |  |  |  |  |  | **3.94(1.070)** |

SD=strongly disagree; D=disagree; N=neutral; A=agree; SA=strongly agree

### Bivariable analysis of organizational strategies and patient waiting time

Bivariable regression analysis established a significant association between sufficient examination rooms (OR 2.24; 95% CI, 1.32-3.77; p-value=0.003, deployment of ushers (OR 2.01; 95% CI, 1.07-3.76; p-value=0.029) and OPD size has got enough space (OR 2.15; 95% CI 1.22-3.79; p-value=0.008) Table 6.

**Table 6:**
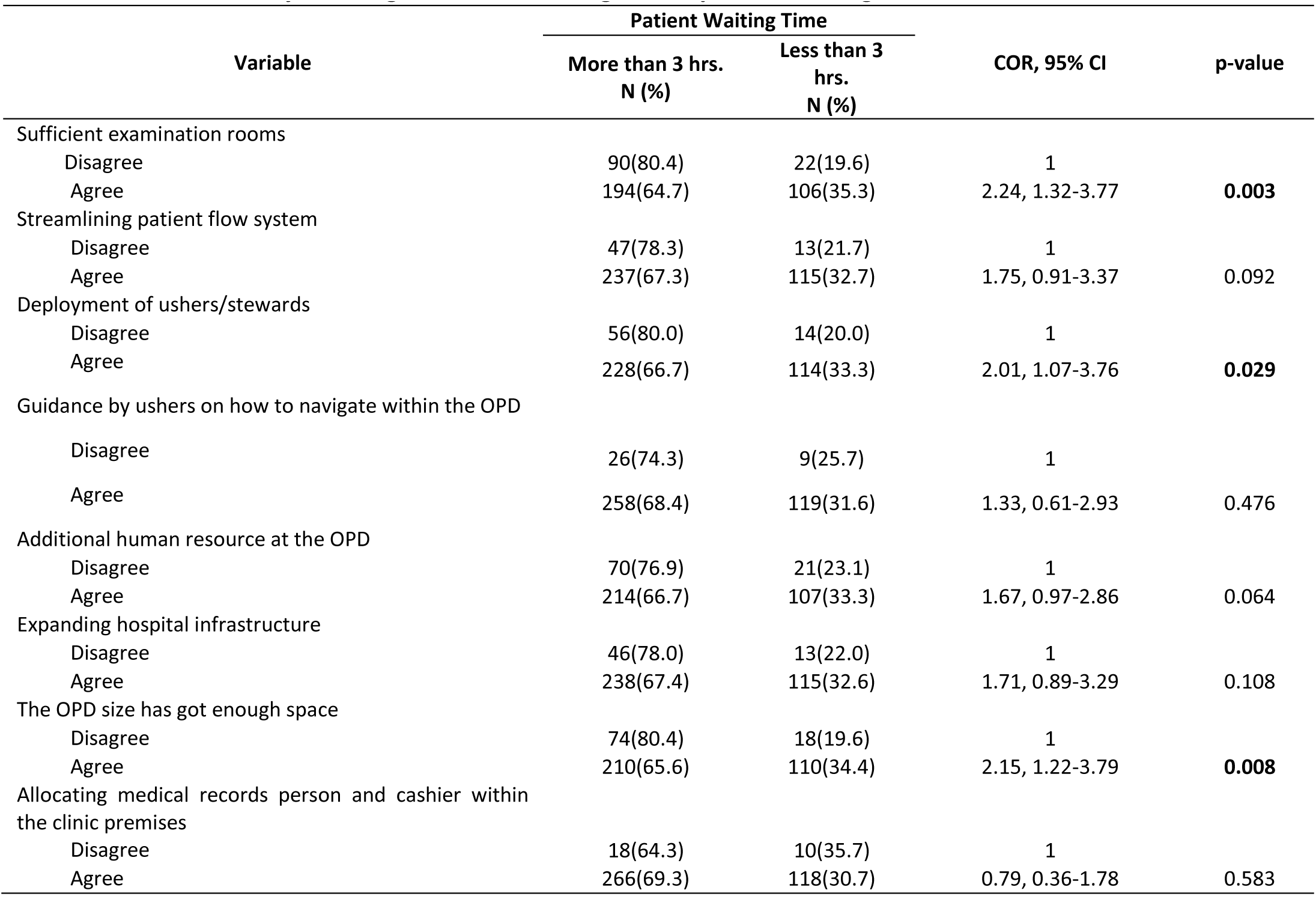
Bivariable analysis of organizational Strategies and patient waiting time.

| Variable | Patient Waiting Time |  | COR, 95% CI | p-value |
| --- | --- | --- | --- | --- |
|  | More than 3 hrs.<br>N (%) | Less than 3<br>hrs.<br>N (%) |  |  |
| Sufficient examination rooms |  |  |  |  |
| Disagree | 90(80.4) | 22(19.6) | 1 |  |
| Agree | 194(64.7) | 106(35.3) | 2.24, 1.32-3.77 | **0.003** |
| Streamlining patient flow system |  |  |  |  |
| Disagree | 47(78.3) | 13(21.7) | 1 |  |
| Agree | 237(67.3) | 115(32.7) | 1.75, 0.91-3.37 | 0.092 |
| Deployment of ushers/stewards |  |  |  |  |
| Disagree | 56(80.0) | 14(20.0) | 1 |  |
| Agree | 228(66.7) | 114(33.3) | 2.01, 1.07-3.76 | **0.029** |
| Guidance by ushers on how to navigate within the OPD |  |  |  |  |
| Disagree | 26(74.3) | 9(25.7) | 1 |  |
| Agree | 258(68.4) | 119(31.6) | 1.33, 0.61-2.93 | 0.476 |
| Additional human resource at the OPD |  |  |  |  |
| Disagree | 70(76.9) | 21(23.1) | 1 |  |
| Agree | 214(66.7) | 107(33.3) | 1.67, 0.97-2.86 | 0.064 |
| Expanding hospital infrastructure |  |  |  |  |
| Disagree | 46(78.0) | 13(22.0) | 1 |  |
| Agree | 238(67.4) | 115(32.6) | 1.71, 0.89-3.29 | 0.108 |
| The OPD size has got enough space |  |  |  |  |
| Disagree | 74(80.4) | 18(19.6) | 1 |  |
| Agree | 210(65.6) | 110(34.4) | 2.15, 1.22-3.79 | **0.008** |
| Allocating medical records person and cashier within the clinic premises |  |  |  |  |
| Disagree | 18(64.3) | 10(35.7) | 1 |  |
| Agree | 266(69.3) | 118(30.7) | 0.79, 0.36-1.78 | 0.583 |

During multivariate logistic analysis the study identified that only deployment of ushers/stewards is significantly associated with reduced patient waiting time with AOR 2.08 (95% CI, 1.10-3.94, p-value=0.025) (Table 7).

**Table 7:** Multivariable analysis of organizational Strategies and patient waiting time.

| Variable | Patient Waiting Time |  | AOR, 95% CI | p-value |
| --- | --- | --- | --- | --- |
|  | More than 3 hrs.<br>N (%) | Less than 3 hrs.<br>N (%) |  |  |
| **Gender** |  |  |  |  |
| Male | 127(66.5) | 64(33.5) | 1 |  |
| Female | 157(71.0) | 64(29.0) | 0.78, 0.51-1.21 | 0.272 |
| **Education** |  |  |  |  |
| Non formal education | 17(85.0) | 3(15.0) | 1 |  |
| Basic education | 202(68.7) | 92(31.3) | 2.44, 0.68-8.74 | 0.170 |
| Collage and above | 65(66.3) | 33(33.7) | 2.75, 0.73-10.29 | 0.134 |
| **Mode of payment** |  |  |  |  |
| Cash | 31(81.6) | 7(18.4) | 1 |  |
| Insurance | 253(67.7) | 121(32.) | 1.64, 0.61-4.38 | 0.324 |
| **There is sufficient examination rooms** |  |  |  |  |
| Disagree | 90(80.4) | 22(19.6) | 1 |  |
| Agree | 194(64.7) | 106(35.3) | 1.97, 0.89-4.37 | 0.096 |
| **Streamlining patient flow system** |  |  |  |  |
| Disagree | 47(78.3) | 13(21.7) | 1 |  |
| Agree | 237(67.3) | 115(32.7) | 0.95, 0.41-2.22 | 0.908 |
| **Deployment of ushers/stewards** |  |  |  |  |
| Disagree | 56(80.0) | 14(20.0) | 1 |  |
| Agree | 228(66.7) | 114(33.3) | 2.08, 1.10-3.94 | **0.025** |
| **Additional human resource** |  |  |  |  |
| Disagree | 70(76.9) | 21(23.1) | 1 |  |
| Agree | 214(66.7) | 107(33.3) | 0.50, 0.18-1.40 | 0.188 |
| **Expanding hospital infrastructure** |  |  |  |  |
| Disagree | 46(78.0) | 13(22.0) | 1 |  |
| Agree | 238(67.4) | 115(32.6) | 1.08, 0.47-2.46 | 0.859 |
| **The OPD size has got enough space** |  |  |  |  |
| Disagree | 74(80.4) | 18(19.6) | 1 |  |
| Agree | 210(65.6) | 110(34.4) | 2.17, 0.81-5.82 | 0.125 |

### Qualitative findings on Organizational strategies Expansion of hospital infrastructure

#### Skepticism with effect on reducing waiting time

The expansion of hospital infrastructure yielded a mix of views from interviewees. Some interviewees expressed skepticism about the efficacy of infrastructure expansion in reducing waiting time, emphasizing that the number of healthcare workers has not increased proportionally. As indicated in quotes below:

> *“Having new numerous buildings alone isn’t sufficient. When we plan to expand hospital infrastructure, it’s imperative to also have a skilled workforce available. Specialized experts are necessary to operate in these newly extended departments”* (IDI – female HCP).

Others viewed that the implementation of hospital infrastructure expansion has yielded positive results in the reduction of patient waiting time in the OPD. A male patient affirmed that:

> *“Being relocated to this new building has made a significant difference. There is minimal congestion, which is tolerable and we are promptly called to see the doctor”* (IDI – male patient).

### Deployment of Ushers

#### Enhanced patient workflow

The implementation of deploying ushers at the OPD has marked a significant stride towards enhancing the overall patient experience and streamlining operations. This proactive step by the management demonstrates a forward-thinking strategy to healthcare service delivery. Their presence not only expedites the process for patients but also contributes to a more efficient and organized workflow within the department. This strategy fosters a positive first impression and sets the tone for the patient’s entire healthcare journey. This is evidenced by unanimous agreement among all participants interviewed and underscores the remarkable effect of deploying ushers in the facility:

> *“The deployment of these ushers by management represents a significant and positive shift. Their function is crucial, not only here but also in highly developed countries where similar roles are assigned. Many of our visitors face difficulties in finding their way, and having these ushers assist them in reaching their destinations is of great importance. While as a doctor, I can give clear instructions to a patient, it presents a different challenge for a visitor. Therefore, the presence of these ushers has played a vital role in speeding up the process for patients to navigate the facility”* (IDI – male HCP).

A female patient said:

> *“When I first arrived, I wasn’t quite sure where to begin. Luckily, as soon as I entered the reception area, I found ushers who promptly directed me to the right starting point. They then informed me about the designated waiting area for doctor consultations, and that saved me more time than I expected”* (IDI – female patient).

Furthermore, another interviewee shared the experience and said:

> “*While walking down the corridor, I found myself uncertain about where to proceed for the investigations. It was then that I met an usher who inquired about my needs and provided clear directions to the specific department. This initial interaction with the usher was instrumental in helping me navigate the facility effectively and get to my destination on time”* (IDI – male patient).

### Additional Human resource

#### Reliance on auxiliary support

The qualitative data reveals a recurring challenge related to staffing levels within the healthcare facility. This was corroborated by a senior staff member who provided insight into the current human resource situation when asked:

> *“In reality, we have encountered staff shortages during different periods. Even now, we rely on the support of volunteers or interns/residents. It’s worth noting that these groups usually serve for a short duration and eventually move on. Nonetheless, their contribution is valuable as it mitigates the shortage of regular staff, even though we haven’t reached the ideal staffing level yet” (IDI – female HCP)*

## Discussion

Following the intervention, it was observed that the overall waiting time in the OPD was reduced to 3.35 hours in contrast to the previous six-hour (6) waiting time prior to the intervention, showing the effectiveness of the intervention achieving a reduction of waiting time by 55.8%. This improvement is significant and suggests that the intervention has had a positive effect, potentially leading to improved waiting time. These findings align with research conducted in hospitals in the USA, China, Sri Lanka, and Taiwan. These hospitals employed various strategies to address long waiting time at the OPD by primarily involving adding more human resources, change in business and management practices. The findings demonstrated a significant success in reducing wait times by 15%, 78%, 60%, and 50%, respectively [27].

The study found that while expanding hospital infrastructure has the potential to reduce waiting times; this relationship did not reach statistical significance, indicating that other factors play a role in waiting time reduction. Qualitative insights underscore the importance of combining infrastructure expansion with a well-trained workforce, emphasizing the need for specialized expertise to efficiently run newly extended departments. Expanding infrastructure is a crucial step in reducing waiting time, but it should be complemented by ensuring the availability of a skilled workforce and considering the overall capacity to accommodate the growing patient population. This finding aligns with previous studies conducted in India [3], Pakistan [5] and Nigeria (Diri and Eledo, 2020) which all highlighted that despite increased demand for healthcare services, medical infrastructure and facilities have not kept pace, resulting in limited working space for medical personnel and patients and contributing to delays in waiting time.

The presence of ushers, responsible for guiding patients in the OPD, has been a significant factor in reducing waiting time, affirming their instrumental role in enhancing the patient experience. Both qualitative and quantitative findings emphasize the indispensable role of ushers in facilitating navigation within the OPD. Their ability to offer clear directions and immediate assistance streamlined the process for patients, contributing to a more efficient and time-effective experience. Patients expressed gratitude for this service, highlighting its contribution to a seamless experience. This role in providing clear communication and guidance significantly expedites patient flow within the facility. These findings are consistency with studies done which were conducted in different countries, which also highlight the value of ushers in reducing waiting times and enhancing patient satisfaction. Studies in the UK [10], India [11], and Haiti [6], as well as in Pakistan [5] and Ethiopia [7], further emphasize the critical role of ushers in expediting patient flow and improving the overall patient experience within healthcare facilities. This positive impact extends beyond reducing waiting time, underlining the importance of integrating such roles into OPD operations for improved patient satisfaction.

Additional human resources at the OPD was perceived positively by respondents, with a mean score of 3.83 (SD=1.203), suggesting a high effect based on the categorization scale used. Moreover, the presence of additional human resources was negatively associated with patient waiting time. In the adjusted odds ratio, the study found that patients who waited for less than 3 hours were significantly less likely to agree that additional human resources have helped to reduce waiting time. In this context, the presence of additional human resources is linked to a significantly lower likelihood of reduction of patient waiting time. Consequently, this suggests that patients continue to perceive a shortage of healthcare workers, despite the allocation of additional resources to address waiting times. These findings are consistent with global studies, including reports from the WHO, which emphasizes the critical shortage of healthcare professionals in many low-income countries [12], including Tanzania. Similar studies in Pakistan [13], Nigeria [14], Kenya [15] and another study that was conducted in rural Tanzania [16] all highlight that a scarcity of healthcare providers contribute to extended waiting time, stressing the need for an adequate workforce to manage patient flow and improve healthcare delivery. The ongoing shortage of human resources for health in Tanzania further stresses the persistent problem of understaffing in the healthcare sector [17]. These collective findings emphasize the pivotal role of an adequate healthcare workforce in reducing patient waiting time and enhancing the quality of care in healthcare facilities.

## Study limitations

Since only one hospital was involved in the study, generalization to cover the rest of Tanzania remains uncertain. In additions, patients were questioned while receiving medical care, which might influence their answers due to the fear of endangering their treatment.

## Conclusion and recommendations

The implemented interventions led to a significant reduction in overall OPD waiting times to an average of 3.30 hours, compared to the previous six-hour wait, demonstrating a 55% efficacy of the implemented strategies. While notable improvements were seen in registration, payment, triage and pharmacy services, challenges persist in waiting for doctor consultations, laboratory procedure and radiology services, leading to extended waiting times for some patients. Organizational strategies, notably the presence of ushers/stewards demonstrated a significant association with reduced waiting time. While expanding hospital infrastructure and increasing human resources proved beneficial, it necessitated additional staffing. Despite these improvements, further enhancements are required to meet the global standard of waiting time ranging from 30 minutes to 2 hours.

## Data Availability

data will be available after acceptance of the manuscript for publication

## Acknowledgments

We extend our gratitude to the patients who participated in this study and the research assistants who contributed to data collection, namely, Geofrey A. Sikaluzwe, Mbayani J. Kivuyo, Richard Hezron Mwamahonje, Emmanuel M. Mabula, Abel E. Lucas, Amos Francis, Dr. (Mrs) Angela Savage for proof reading and Dr. Bernard Njau.

## Author contributions

**Conceptualization:** Manasseh Joel Mwanswila

**Formal analysis:** Manasseh Joel Mwanswila

**Investigation:** Manasseh Joel Mwanswila

**Methodology:** Manasseh Joel Mwanswila, Henry Mollel

**Supervision:** Henry Mollel

**Validation:** Henry Mollel

**Writing – original draft:** Manasseh Joel Mwanswila

**Writing – review & editing:** Henry Mollel

